# Risk factors for mortality among hospitalized patients with COVID-19

**DOI:** 10.1101/2020.09.22.20196204

**Authors:** Devin Incerti, Shemra Rizzo, Xiao Li, Lisa Lindsay, Vince Yau, Dan Keebler, Jenny Chia, Larry Tsai

**Author notes:** **Correspondence to:** Devin Incerti, PhD.

## Abstract

**Objectives:** To develop a prognostic model to identify and quantify risk factors for mortality among patients admitted to the hospital with COVID-19.

**Design:** Retrospective cohort study. Patients were randomly assigned to either training (80%) or test (20%) sets. The training set was used to fit a multivariable logistic regression. Predictors were ranked using variable importance metrics. Models were assessed by C-indices, Brier scores, and calibration plots in the test set.

**Setting:** Optum® de-identified COVID-19 Electronic Health Record dataset.

**Participants:** 17,086 patients hospitalized with COVID-19 between February 20, 2020 and June 5, 2020.

**Main outcome measure:** All-cause mortality during hospital stay.

**Results:** The full model that included information on demographics, comorbidities, laboratory results and vital signs had good discrimination (C-index = 0.87) and was well calibrated, with some overpredictions for the most at-risk patients. Results were generally similar on the training and test sets, suggesting that there was little overfitting.

Age was the most important risk factor. The performance of models that included all demographics and comorbidities (C-index = 0.79) was only slightly better than a model that only included age (C-index = 0.76). Across the study period, predicted mortality was 1.2% for 18-year olds, 8.4% for 55-year olds, and 28.6% for 85-year olds. Predicted mortality across all ages declined over the study period from 21.7% by March to 13.3% by May.

**Conclusion:** Age was the most important predictor of all-cause mortality although vital signs and laboratory results added considerable prognostic information with oxygen saturation, temperature, respiratory rate, lactate dehydrogenase, and white blood cell count being among the most important predictors. Demographic and comorbidity factors did not improve model performance appreciably. The model had good discrimination and was reasonably well calibrated, suggesting that it may be useful for assessment of prognosis.

## Introduction

In December 2019 an outbreak of novel coronavirus disease 2019 (COVID-19) occurred in Wuhan, China, and was officially declared a pandemic in March 2020 by the World Health Organization (WHO). Severe COVID-19 illness can result in hospitalization, intensive care stays, or even death. To date (September 2020), more than 27 million people have been infected worldwide and over 900,000 people have died [1]. Mortality rates among hospitalized patients are especially high and have ranged from 15 to 20 percent in the United States (U.S.) [2-4].

High mortality rates are due to a number of factors including severity of the disease, a lack of available treatments, and in some cases shortages in medical supplies and personnel caused by surges in hospitalizations. Yet, despite high mortality rates, there is still considerable uncertainty about who is at most risk for the worst outcomes. Efforts to reduce this uncertainty through better quantification of the relative importance of risk factors for severe illness can help on a number of fronts. For one, it has been suggested that known risk factors for severe outcomes can be used by medical staff to triage patients or for health systems to identify priority groups for vaccination [5-7]. Furthermore, it can help individuals understand their own risks of illness and clinicians assess prognosis for their patients. Finally, it can be used to stratify patients in clinical trials or to identify covariates to adjust for in comparative-effectiveness analyses using observational data [8-10].

For many of these reasons there has been a strong interest in the development of prognostic models for COVID-19. While a number of models have already been developed, there is still a need for more rigorous and validated models. Wynants et al. [5] conducted a systematic review of existing models and found that most studies had a high likelihood of bias due to non-representative patient cohorts, overfitting due to small sample sizes, and exclusion of patients who have not yet had an event. Similarly, studies have inadequately adjusted for confounding variables when assessing the impact of particular risk factors. It is therefore difficult to make claims on the relative importance of different risk factors [11].

In this study, we developed a prognostic model of in-hospital mortality and aimed to overcome many of the limitations of prior studies. Our model was trained using a large sample of 13,658 hospitalized patients with COVID-19 from an Electronic Health Record (EHR) Database encompassing all regions of the U.S. Multivariable regression techniques were used to isolate the independent impact of risk factors and quantify their relative importance. Validation was performed by assessing the model’s out-of-sample predictive performance, including on a large test dataset of 3,428 patients not used in model fitting. To facilitate transparency in research, all code for the analysis is available at https://github.com/phcanalytics/covid19-prognostic-model and included in the Supplement.

## Methods

### Data Source

The Optum® de-identified COVID-19 EHR dataset was used to identify patients hospitalized with COVID-19. The dataset consists of a national sample of inpatient and outpatient medical records sourced from hospital networks from across the U.S. Data are de-identified in compliance with the HIPAA Expert Method and managed according to Optum customer data use agreements [4]. Inpatient mortality was captured in the dataset as month of death. Age for those 89 and older was aggregated in the dataset.

### Study Cohort

To be eligible for the hospitalized cohort in this study, patients were required to be older than 18 years old and have: (1) a U07.1 or U07.2 diagnosis, (2) a positive SARS-CoV-2 diagnostic test (e.g., either molecular or antigen tests) or (3) a B97.29 diagnosis with the absence of a negative SARS-CoV-2 molecular test within a 14-day window. Eligible hospitalizations required inpatient or emergency room overnight visits with a COVID-19 diagnosis or starting up to 21 days after a COVID-19 diagnosis. Contiguous ER and inpatient visits, with up to a 1-day gap were considered a single hospitalization. The date of admission was used as index date when COVID-19 diagnosis occurred before hospitalization, otherwise index date was set to the date of COVID-19 diagnosis. Only the first hospitalization was considered in this study. The study period was February 20, 2020 to June 5, 2020.

### Outcome

The outcome was a binary measure of in-hospital all-cause mortality. To ensure that there was sufficient follow-up time, patients were removed from the sample if their index date was less than 2 weeks prior to the date the Optum dataset was censored (June 5), resulting in the removal of approximately 5% of the overall study population. Evidence from the U.S. Centers for Disease Control on time to death following hospital admission (median: 5 days, interquartile-range: 3-8 days) [12] suggests that this was sufficient to capture most deaths.

### Predictors

Candidate predictors were chosen based on prior research and/or clinical consultation [5]. They included demographics, comorbidities, vital signs, laboratory results and calendar time. Demographic variables were age, sex, race, ethnicity, geographic division, and smoking status. Comorbidities were identified based on ICD-9 and ICD-10 codes within a year of index date and included hypertension, diabetes, and those included in the Charlson Comorbidity Index (CCI).

Vital signs considered were peripheral oxygen saturation, systolic blood pressure, heart rate, respiratory rate, temperature, and body mass index (BMI). Laboratory results for aspartate aminotransferase (AST), C-reactive protein (CRP), creatinine, ferritin, lactate dehydrogenase (LDH), troponin I, lymphocyte count, neutrophil count, platelet count (PLT), and white blood cell count (WBC) were obtained. D-dimer and procalcitonin laboratory measures were also considered but were dropped due to very high missingness (90% and 49%, respectively). We restricted vital signs and laboratory results to those within a [-3,+1] day window surrounding the index date and used the (median of the) value(s) closest to the index date when multiple values were available within the window.

To capture trends over time, we also derived a calendar time variable measured as the number of days between a patient’s index date and the date of the first case in the data.

Missing data for each of the candidate predictors is summarized in **Supplement Section 3**.**2**. There was significant missing data for race, ethnicity, BMI, smoking status, and some of the vital signs and laboratory results. Multiple imputation was consequently used as described below.

### Model Development and Statistical Analysis

A multivariable logistic regression was used to model mortality. To protect against overfitting, we first performed variable selection by fitting a logistic model with a group lasso penalty [13,14]. We repeatedly fit the lasso model 100 times and used 10-fold cross-validation during each of the 100 iterations to select the optimal tuning parameter, “lambda.” Variables with non-zero coefficients in at least 90 percent of the iterations were included. In practice, only two variables were excluded: peptic ulcer disease and neutrophil count (**Supplement Section 5**).

The model with all predictors chosen by group lasso was the “full” model. For comparison, we fit 4 more parsimonious models with the following predictors: (i) age only, (ii) comorbidities only, (iii) all demographics (and calendar time), and (iv) demographics (and calendar time) and comorbidities. Non-linear relationships between mortality and continuous covariates were modeled using restricted cubic splines. 3 knots were deemed sufficient based on graphical assessment of univariate fits in nearly all cases; 4 knots were used for respiration rate to capture a bathtub-shaped relationship with mortality. These graphical assessments also showed that a few outliers were influencing the fit of some of the laboratory results. They were consequently truncated from above (at the “outer fence” defined as the third quartile plus three times the interquartile range), which led to stronger and more clinically meaningful relationships (**Supplement Section 3**.**5)**.

Predictor effects were summarized in four ways. First, coefficient estimates were translated into clinically meaningful odds ratios. Odds ratios for each value of a categorical variable were computed based on comparisons to a reference group. Odds ratios for continuous covariates were based on changes in value from the 25th percentile to the 75th percentile. Second, we predicted log odds across different values of each predictor and visually assessed the effects. Third, predicted probabilities were computed by age, sex, and calendar time for a random sample of 1,000 patients and then averaged across patients. Fourth, variable importance was assessed using □2 minus degrees of freedom from a Wald test that tests the hypothesis that the coefficient of each term associated with a variable (e.g., all categories of a categorical variable or all spline terms of a continuous variable) is zero [15,16].

To validate the model, we randomly split the data into a training and test set using an 80/20 split and evaluated the model in both the training and the test sets. Model performance was assessed using the C-index (AUROC) and Brier score. To assess overfitting, 50 bootstrap replications were used to quantify “optimism” in the training set, defined as the average of differences in model performance between the training and bootstrapped samples. Calibration was assessed using a calibration curve that compared predicted probabilities with actual probabilities.

Missing data was imputed using multivariate imputation by chained equations (MICE) [18]. A total of 5 datasets were imputed. Assessment of the imputation was performed by comparing the distribution of the missing imputed data to the observed data for each predictor. **Supplement Section 4**.**3** shows that these distributions were very similar for each variable, which suggests that the imputation was adequate. Coefficient estimates and confidence intervals were combined across the imputations using Rubin’s rule [17].

Analyses were performed using R 4.0.0. We used three main R packages in the analysis: *mice* for multiple imputation, *oem* to fit the group lasso, and *rms* to fit the multivariable logistic regressions, summarize the coefficients, and validate the model [14,15,18]. Additional details of the methodology are available in the Supplement.

## Results

### Characteristics of the Study Population

We identified 17,086 patients that met the inclusion criteria for the COVID-19 hospitalized cohort. The characteristics of the 13,658 patients included in the training set are described in **Table 1** and in additional detail in **Supplement Section 3**.

**Table 1.** Characteristics of hospitalized patients with COVID-19 in training set by mortality status.

|  | Missing (%) | Overall<br>Median (IQR)<br>N (%) | Survivor<br>Median (IQR)<br>N (%) | Non-survivor<br>Median (IQR)<br>N (%) |
| --- | --- | --- | --- | --- |
| **N** |  | 13658 | 11495 | 2163 |
| **Demographics** |  |  |  |  |
| Age, median (IQR) | 0 | 62.0 [49.0, 75.0] | 59.0 [46.0, 71.0] | 77.0 [67.0, 85.0] |
| Calendar time (days), median (IQR) | 0 | 47.0 [38.0, 64.0] | 47.0 [38.0, 64.0] | 46.0 [37.0, 60.0] |
| Geographic division (%) | 2.7 |  |  |  |
| East North Central |  | 4627 (34.8) | 3954 (35.3) | 673 (31.9) |
| Middle Atlantic |  | 4636 (34.9) | 3844 (34.4) | 792 (37.6) |
| New England |  | 1583 (11.9) | 1272 (11.4) | 311 (14.8) |
| Other |  | 191 ( 1.4) | 175 ( 1.6) | 16 ( 0.8) |
| Pacific |  | 511 ( 3.8) | 438 ( 3.9) | 73 ( 3.5) |
| South Atlantic/West South Central |  | 364 ( 2.7) | 317 ( 2.8) | 47 ( 2.2) |
| West North Central |  | 1384 (10.4) | 1189 (10.6) | 195 ( 9.3) |
| Race / Ethnicity | 26.1 |  |  |  |
| Non-Hispanic white |  | 5647 (56.0) | 4455 (53.1) | 1192 (70.3) |
| Asian |  | 362 ( 3.6) | 307 ( 3.7) | 55 ( 3.2) |
| Hispanic |  | 533 ( 5.3) | 478 ( 5.7) | 55 ( 3.2) |
| Non-Hispanic black |  | 3547 (35.2) | 3153 (37.6) | 394 (23.2) |
| Sex = Female/Male (%) | 0 | 6563/7091<br>(48.1/51.9) | 5635/5856 (49.0/51.0) | 928/1235<br>(42.9/57.1) |
| Smoking status (%) | 25.6 |  |  |  |
| Current |  | 866 ( 8.5) | 785 ( 9.0) | 81 ( 5.5) |
| Never |  | 6207 (61.1) | 5450 (62.8) | 757 (51.1) |
| Previous |  | 3092 (30.4) | 2450 (28.2) | 642 (43.4) |
| **Comorbidities** |  |  |  |  |
| Acute myocardial infarction | 0 | 1535 (11.2) | 1028 ( 8.9) | 507 (23.4) |
| AIDS/HIV | 0 | 101 ( 0.7) | 89 ( 0.8) | 12 ( 0.6) |
| Cancer | 0 | 1678 (12.3) | 1282 (11.2) | 396 (18.3) |
| Cerebrovascular disease | 0 | 1439 (10.5) | 1023 ( 8.9) | 416 (19.2) |
| Congestive heart failure | 0 | 3627 (26.6) | 2933 (25.5) | 694 (32.1) |
| Chronic pulmonary disease | 0 | 2325 (17.0) | 1604 (14.0) | 721 (33.3) |
| Dementia | 0 | 1394 (10.2) | 854 ( 7.4) | 540 (25.0) |
| Diabetes | 0 | 4612 (33.8) | 3669 (31.9) | 943 (43.6) |
| Hemiplegia or paraplegia | 0 | 330 ( 2.4) | 228 ( 2.0) | 102 ( 4.7) |
| Hypertension | 0 | 8003 (58.6) | 6333 (55.1) | 1670 (77.2) |
| Metastatic cancer | 0 | 277 ( 2.0) | 188 ( 1.6) | 89 ( 4.1) |
| Mild liver disease | 0 | 879 ( 6.4) | 711 ( 6.2) | 168 ( 7.8) |
| Moderate/severe liver disease | 0 | 128 ( 0.9) | 88 ( 0.8) | 40 ( 1.8) |
| Peptic ulcer disease | 0 | 206 ( 1.5) | 160 ( 1.4) | 46 ( 2.1) |
| Peripheral vascular disease | 0 | 1671 (12.2) | 1176 (10.2) | 495 (22.9) |
| Renal disease | 0 | 2833 (20.7) | 1984 (17.3) | 849 (39.3) |
| Rheumatoid disease | 0 | 398 ( 2.9) | 315 ( 2.7) | 83 ( 3.8) |
| CCI | 0 | 1.0 [0.0, 3.0] | 1.0 [0.0, 2.0] | 3.0 [1.0, 5.0] |
| **Vitals** |  |  |  |  |
| BMI, median (IQR) | 11.9 | 29.7 [25.5, 35.1] | 30.0 [25.8, 35.4] | 28.1 [24.0, 33.5] |
| Diastolic blood pressure (mm Hg), median (IQR) | 3.1 | 73.0 [65.5, 80.5] | 74.0 [66.5, 81.0] | 68.0 [60.0, 75.5] |
| Heart rate (beats/min), median (IQR) | 3.1 | 87.5 [77.5, 98.0] | 87.0 [77.5, 98.0] | 89.0 [77.5, 102.0] |
| Oxygen saturation (%), median (IQR) | 3.9 | 96.0 [94.0, 98.0] | 96.0 [94.5, 98.0] | 95.0 [93.0, 97.0] |
| Respiratory rate (breaths/min), median (IQR) | 3.9 | 20.0 [18.0, 22.0] | 19.5 [18.0, 21.0] | 22.0 [19.0, 26.0] |
| Systolic blood pressure (mm Hg), median (IQR) | 3.2 | 126.0 [115.0, 139.0] | 127.0 [116.0, 139.0] | 122.0 [109.0, 136.5] |
| Temperature (Celsius), median (IQR) | 3.1 | 37.0 [36.7, 37.4] | 37.0 [36.7, 37.4] | 37.1 [36.7, 37.6] |
| **Laboratory tests** |  |  |  |  |
| Alanine aminotransferase (ALT) (U/L), median (IQR) | 20.1 | 28.0 [18.0, 46.0] | 28.0 [18.0, 46.0] | 27.0 [18.0, 44.0] |
| Aspartate aminotransferase (AST) (U/L), median (IQR) | 21 | 37.0 [25.0, 58.0] | 35.0 [25.0, 54.0] | 46.0 [30.0, 73.0] |
| C-reactive protein (mg/L), median (IQR) | 38.7 | 79.1 [34.0, 140.0] | 72.2 [30.0, 130.0] | 116.0 [63.0, 184.0] |
| Creatinine (mg/dL), median (IQR) | 10.4 | 1.0 [0.8, 1.4] | 1.0 [0.8, 1.3] | 1.3 [1.0, 2.1] |
| Ferritin (ng/mL), median (IQR) | 43.6 | 510.0 [224.0, 1080.0] | 470.0 [207.0, 992.0] | 747.5 [320.8, 1501.5] |
| Fibrin D-Dimer (ng/mL), median (IQR) | 90.4 | 750.0 [390.0, 1540.8] | 692.5 [370.0, 1346.5] | 1345.0 [668.2, 3315.0] |
| Lactate dehydrogenase (LDH) (U/L), median (IQR) | 45.2 | 321.0 [238.0, 441.0] | 308.0 [232.0, 415.0] | 404.0 [284.0, 556.5] |
| Lymphocyte count ($10^3/\mu\text{L}$), median (IQR) | 11.2 | 1.0 [0.7, 1.4] | 1.0 [0.7, 1.4] | 0.8 [0.5, 1.1] |
| Neutrophil count ($10^3/\mu\text{L}$), median (IQR) | 11.2 | 4.9 [3.4, 7.1] | 4.7 [3.2, 6.7] | 6.1 [4.1, 9.2] |
| Platelet count (PLT) ($10^3/\mu\text{L}$), median (IQR) | 9.8 | 202.0 [157.0, 260.0] | 205.0 [160.0, 262.0] | 187.5 [143.0, 245.0] |
| Procalcitonin (ng/mL), median (IQR) | 49.3 | 0.1 [0.1, 0.4] | 0.1 [0.1, 0.3] | 0.3 [0.1, 1.0] |
| Troponin (ng/mL), median (IQR) | 41.2 | 0.0 [0.0, 0.0] | 0.0 [0.0, 0.0] | 0.0 [0.0, 0.1] |
| White blood cell count (WBC) ($10^3/\mu\text{L}$), median (IQR) | 9.7 | 6.7 [4.9, 9.1] | 6.5 [4.8, 8.7] | 7.7 [5.6, 11.1] |

The median age was 62 years old (Interquartile range [IQR]: 49, 75). The cohort was composed mostly of male (51.9%) non-Hispanic whites (56.0%). Most patients resided in the Middle Atlantic (34.9%) and East North Central (34.9%) geographic divisions, which mirrors the initial surge of cases in the U.S. Patients had high rates of comorbidities: 58.6% had hypertension, 33.8% had diabetes, 26.6% had chronic pulmonary disease (CPD), 20.7% had renal disease, and the median CCI score was 1 (IQR: 0, 3). The majority of patients were overweight (30%) or obese (48%). Median oxygen saturation was 96.0%, and 25% of patients had oxygen saturation lower than 94.0%. The median temperature was 37.0 C (IQR: 36.7, 37.4). Patients had a median ferritin of 510.0 ng/ml (IQR: 224.0, 1080.0) and median CRP of 79.1 mg/l (IQR: 34.0, 140.0). Among the 9.6% of patients with results available for fibrin d-dimer, the median result was 750 ng/ml (IQR: 390.0, 1540.8).

Compared to survivors, non-survivors were more likely to be male (57.1% vs. 51.0%) and older (median 77 years old vs. 59 years old). Non-survivors also had more comorbidities (median CCI of 3 [IQR: 1,5] vs. 1 [IQR: 0, 2]) including dementia (25.0% vs. 7.4%), renal disease (39.3% vs 17.3%), diabetes (43.6% vs 31.9%), and hypertension (77.2% vs. 55.1%). Similar trends were observed for laboratory findings and vitals: non-survivors had higher heart rates (median beats/min of 89.0 vs. 87.0) and respiratory rates (median breaths/min of 22.0 vs. 19.5), and lower oxygen saturation (median 95.0% vs. 96.0%). However, no substantial difference was observed in temperature (median 37.1 C vs 37.0 C) or in BMI (median 28.1 vs. 30.0). Non-survivors had higher CRP (median mg/L of 116.0 vs 72.2), ferritin (median ng/mL 747.5 vs. 470.0), and fibrin d-dimer (median ng/mL of 1,345 vs 692.5).

A comparison of the training and test sets is provided in **Supplement Section 8**.**1**. There were no meaningful differences in demographics, comorbidities, vitals or laboratory results.

### Predictor Effects

Odds ratios from the multivariable logistic regression are displayed in **Figure 1**. Age was a very important predictor as the odds of death for a 75 year old (75th percentile) were around 6 times than for a 49 year old (25th percentile). Metastatic cancer, moderate/severe liver disease, hemiplegia/paraplegia, and dementia were also highly positively associated with mortality and precisely estimated. Mortality decreased over time as evidenced by the negative odds ratio for calendar time. Other important predictors were vital signs and laboratory results. Lower values of PLT and higher values of AST, troponin I, CRP, WBC, LDH, and creatinine were associated with increased mortality. Similarly, abnormally low levels of oxygen saturation were associated with higher mortality as were abnormally high values of respiration rate, heart rate, temperature, and BMI. Since some of the predictor effects were non-linear, plots showing the non-linear predicted effects of the continuous variables on the log odds scale are displayed in the **Supplement Section 6**.**6**. The log-odds plot shows, for instance, that mortality is higher at both lower and higher levels of temperature and systolic blood pressure, and that the strong negative relationship between oxygen saturation and mortality is only present below around 95%.

**Figure 1:**
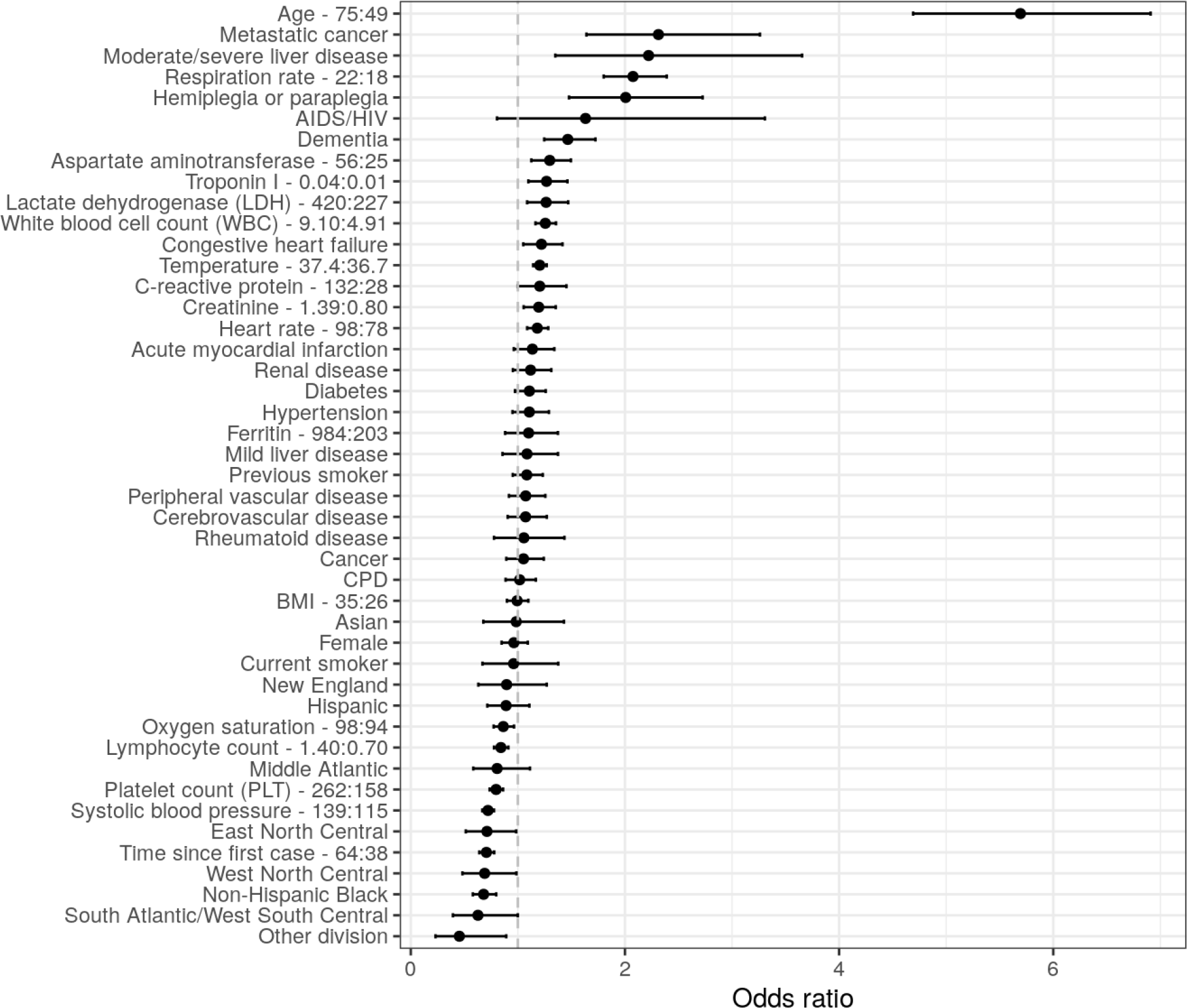
Odds ratios of mortality from the full multivariable logistic regression. Error bars represent 95% confidence levels. Interquartile-range odds ratios are used for continuous predictors (upper quartile: lower quartile). Reference groups for categorical predictors are as follows: race/ethnicity = “non-Hispanic white”, division = “Pacific”, sex = “Male”, smoking = “Never smoker”.

One limitation of the odds ratio is that it can be difficult to compare the relative importance of categorical and continuous variables or transformed continuous variables. **Figure 2** consequently displays the importance of each variable based on Wald tests. Age is the most important predictor by a considerable amount. Laboratory results and vitals tend to be more important predictors than either comorbidities or demographics: impactful predictors include respiration rate, temperature, oxygen saturation, heart rate, WBC, and LDH. Calendar time is also an important predictor.

**Figure 2:**
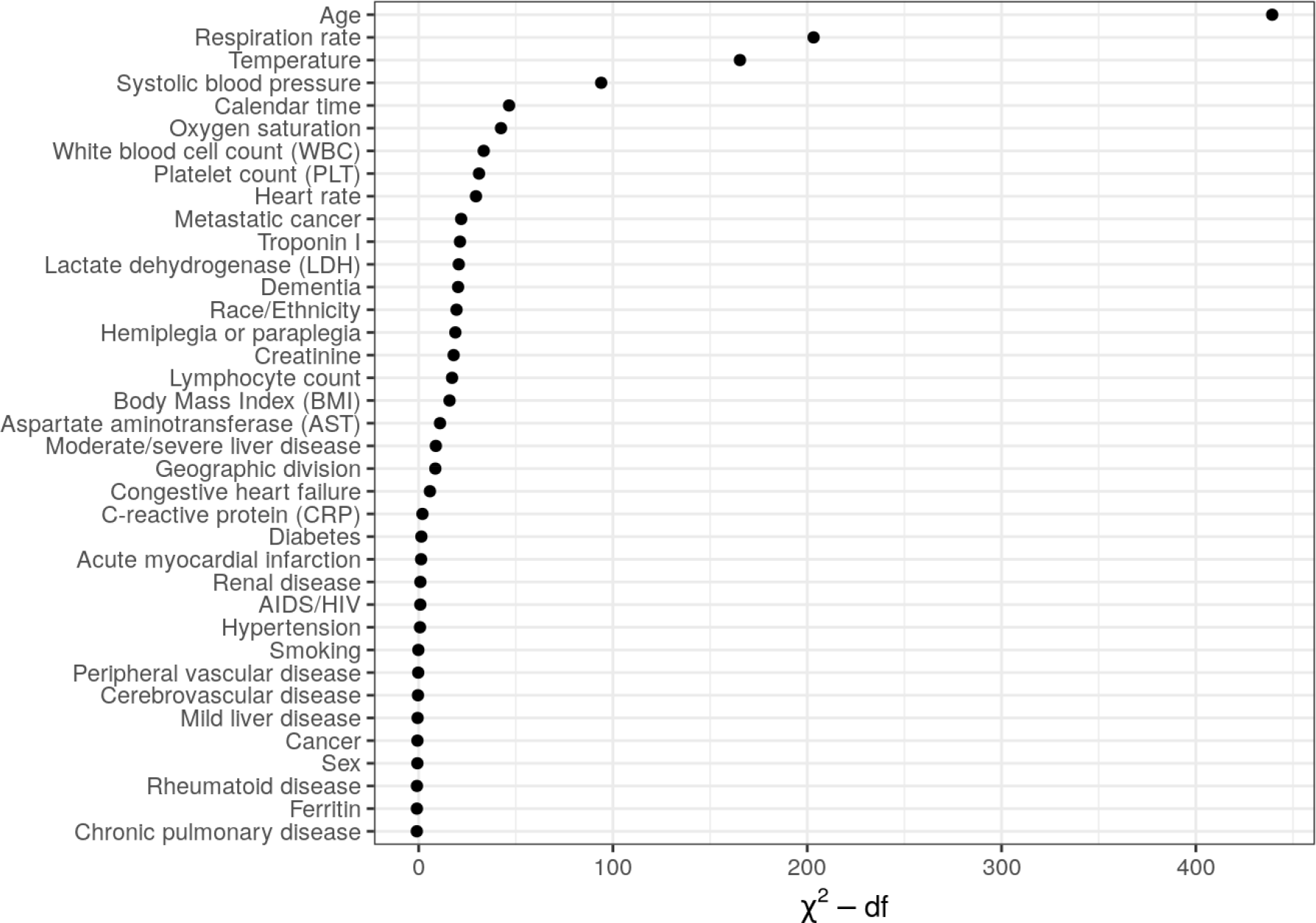
Ranking of importance of predictors of mortality from the full multivariable logistic regression. A higher value of “Chi-squared minus degrees of freedom” implies that a predictor has a larger contribution to the fit of the model.

One potential concern with these results is that laboratory results and vitals might be mediators that are correlated with demographics and comorbidities (**Supplement Section 3**.**4**.**4)**. We assessed this in **Supplement Section 6**.**5** by removing labs and vitals from the model. The odds ratios tended to be fairly stable. Female sex was a notable exception as the odds ratio changed from approximately 0.7 to almost 1.

To assess the clinical impact of changes in predictors, it is helpful to assess predicted probabilities of mortality. These are displayed across levels of age and calendar time for a random sample of 1,000 patients in **Figure 3**. The effect of age on the probability scale is exponential and increasingly sharply at older ages. In March of 2020, a hospitalized 80-year-old had a predicted probability of death of 34% whereas a 70-year-old had a predicted probability of death around 10 percentage points lower. The predicted probability of death for an 18-year-old at the same date was less than 2%. Mortality probabilities did decrease considerably over time with mortality rates for 80-year-olds approaching 20% by May 2020.

**Figure 3:**
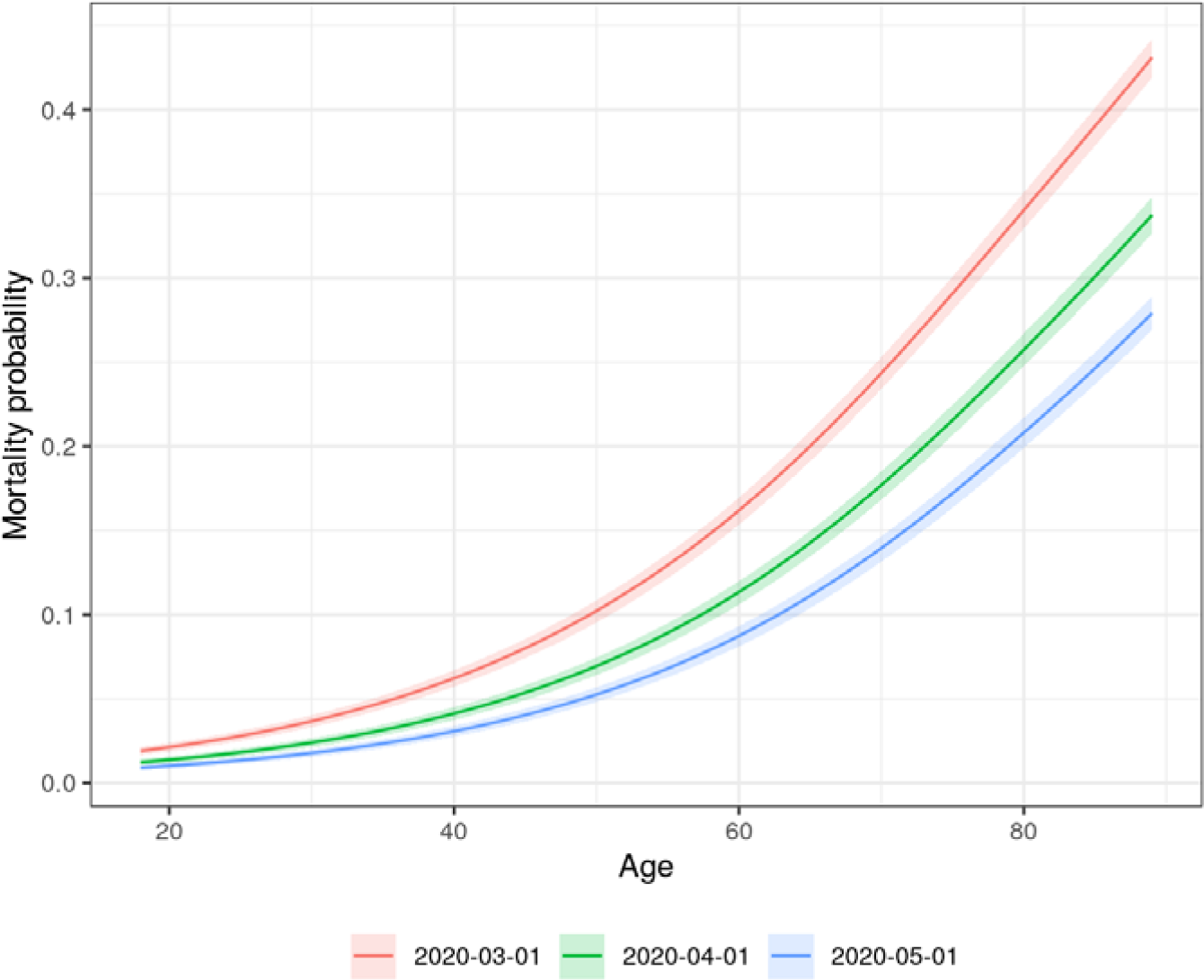
Predicted probability of mortality from the full multivariable logistic regression by age and calendar time. Each curve represents a specific hypothetical index date. Age and calendar time effects are adjusted for all variables in the full model. Predictions for each age and calendar time combination are averaged over a random sample of 1000 patients.

### Validation and Predictive Performance

Calibration curves comparing predictive probabilities to actual probabilities among patients in the training and test sets are shown in **Supplement Section 7**.**2** and **Figure 4**, respectively. In the training set, the bias-adjusted and unadjusted curves were similar, and “optimism” was approximately zero, suggesting that there was little overfitting; however, there was more overprediction for higher risk patients in the test set. In both cases, models including comorbidities tended to be poorly calibrated for more at risk patients, while the full model was better calibrated.

**Figure 4:**
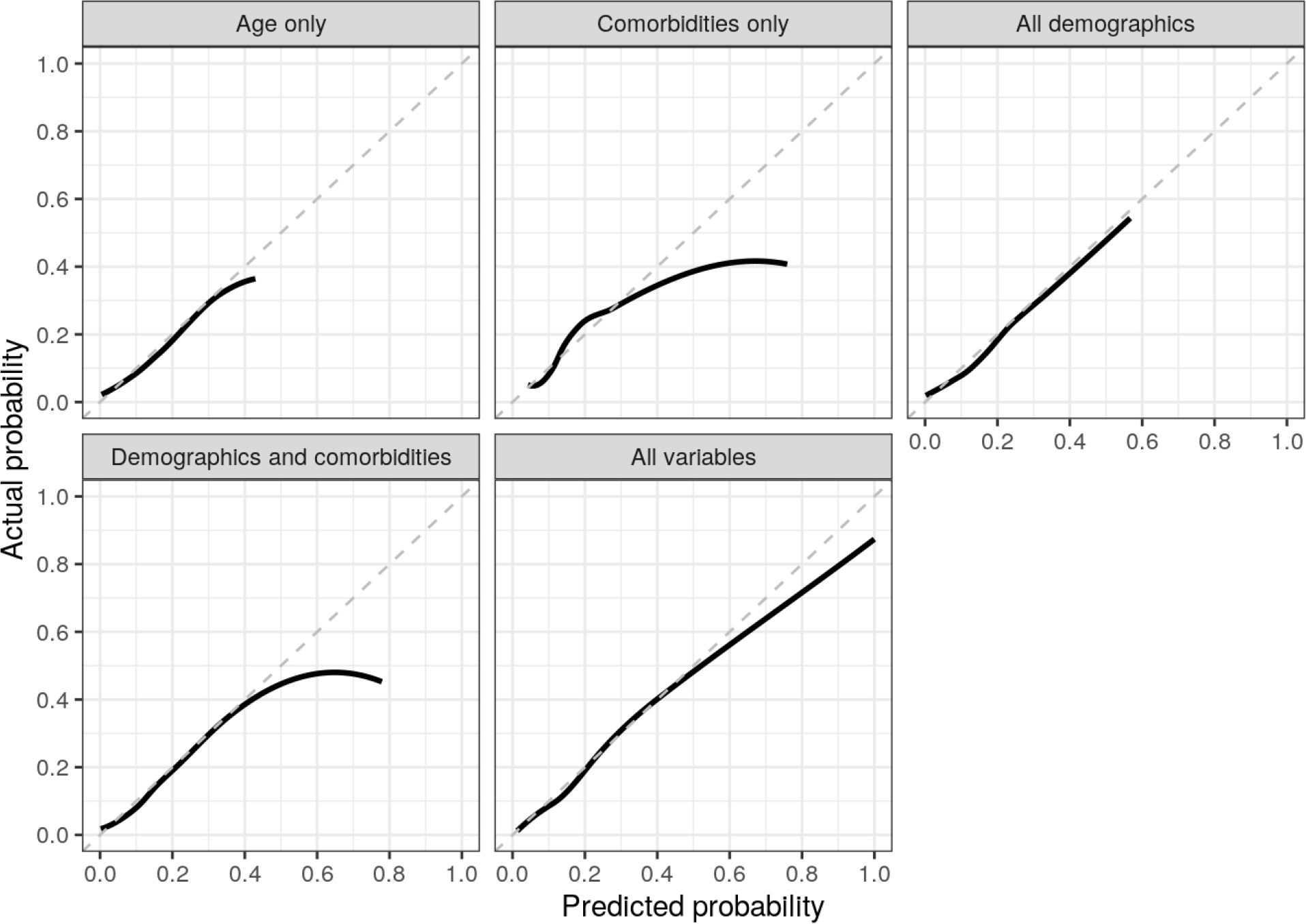
Calibration curves from predictions of the logistic regression model on the test set by model specification. Points on the dashed 45-degree line imply that the predicted probability is equal to the actual probability.

Higher predicted probabilities of mortality were much more common in the full model than in any of the other models. In other words, the variance of the predictions was higher implying that the full model was better able to discriminate between patients. This was also reflected in **Table 2**, which reported estimates of the C-index and Brier score. In the test set, the C-index improved from 0.756 in the age only model to 0.874 in the full model and the Brier score decreased from 0.111 to 0.087. Results were similar in the training set. Notably, there was no appreciable difference in either the Brier score or the C-index between the age only model, the demographics model, or the demographics and comorbidities model.

**Table 2:** Summary of predictive performance in the training and test sets by model specification.

|  | Training set |  | Test set |  |
| --- | --- | --- | --- | --- |
| **Model** | **C-index (AUROC)** | **Brier score** | **C-index (AUROC)** | **Brier score** |
| Age only | 0.7746 | 0.1159 | 0.7558 | 0.1111 |
| All comorbidities | 0.7310 | 0.1216 | 0.7186 | 0.1151 |
| All demographics | 0.7841 | 0.1145 | 0.7732 | 0.1083 |
| Demographics and comorbidities | 0.8014 | 0.1119 | 0.7904 | 0.1062 |
| All variables | 0.8822 | 0.0897 | 0.8741 | 0.0877 |

## Discussion

We aimed to develop a rigorous model that was both clinically interpretable and had good predictive performance. We quantified the relative importance of different predictors to identify risk factors that would be most important for assessing prognosis. One of the most striking findings from our study is that the most parsimonious model---one that only includes age---is highly predictive; in fact, age is nearly as prognostic as all other information on demographic and comorbidities. This does not mean that age alone is sufficient for prediction, but it does suggest that simply knowing a patient’s age is very informative. Vital signs and laboratory results do improve model predictions which use age alone, meaningfully increasing the C-index and decreasing the Brier score.

### Comparison with Other Studies

A living systematic review of existing prognostic models has been conducted by Wynants et al. and 50 prognostic models have been identified to date. A subset of those predict mortality among hospitalized patients.

Most prognostic models are based on data from China [19-24], although others have been developed with data from the United Kingdom [25,26], Mexico [27], South Korea [28], Israel [29], the U.S. [30-32]), and a mix of countries [33]. Our study differs from the other U.S. studies in that it includes a broader cohort of patients encompassing all geographic regions.

Models have typically been trained on smaller datasets, with most consisting of <1,000 and a few ranging from 1000-3000 [28,30,31]. Only one study, to our knowledge, had a sample size larger than 10,000 patients and was based in Mexico [27]. Smaller sample sizes not only increase the likelihood of overfitting, but also reduce the ability to detect important risk factors.

One advantage of our approach compared to more “black box” prediction models, is that the effects of the covariates were interpretable [34,35]. For instance, we used splines [17,18,36] to show that the probability of death increases exponentially with age. The strength of this relationship was consistent with other studies including Gupta et al. [26], who evaluated the performance of models from 19 studies (14 of which were COVID-19 specific) on an independent dataset of 411 patients hospitalized for COVID-19 in the United Kingdom and found that no other variables added additional incremental value beyond age. Age may be a strong predictor because while it is correlated with the comorbidities specified in the model, it also likely captures other unspecified comorbidities that may be associated with worse outcomes. Additionally, age may be associated with altered immune function that could result in slower viral clearance, or a hyperactive immune response that could contribute to severe clinical manifestations of the disease [37].

Our results also differ from Gupta et al. [26] in that laboratory results and vital signs added non-negligible incremental value to the discriminative ability of our predictions. Oxygen saturation, temperature, and respiration rate were among the most important risk factors in the model, which is consistent with other models of mortality, both amongst patients with COVID-19 [22,25,38] and in more general populations [39-41]. The positive relationship between higher levels of BMI and mortality was consistent with prior research [42,43].

Troponin I, LDH, and PLT were among the most important laboratory results, which has been documented elsewhere [5,44,45]. Troponin and LDH elevations may reflect more severe microvascular dysfunction which could lead to myocardial and other end-organ injury. Lower PLT counts could reflect increased consumption due to macro- and micro-thromboses, which have been described clinically and in autopsy studies [46], and which may be associated with and exacerbate microvascular dysfunction. Of note, the lasso model selected WBC and lymphocyte count for inclusion and excluded neutrophil count. We explored this further by running a separate model omitting WBC and found that neutrophil count had similar variable importance to WBC in this specification. Neutrophil count also had a strong relationship with mortality in univariate fits (**Supplement Section 3**.**5)**. Our results are therefore consistent with prior work showing that mortality is associated with lower lymphocyte and higher neutrophil counts [47,48]. We did not find an association between ferritin and mortality despite studies showing that severe illness is characterized by hyperferritinemia [49].

Finally, while comorbidities added little prognostic information beyond age, it is important to distinguish these findings from those based on a general population diagnosed with COVID-19 since risk factors that are predictive of hospitalization (or death in the general population) may not be predictive of mortality conditional on hospitalization. Petrilli et al. [44] provides some evidence consistent with this in that comorbidities were more important predictors of hospital admission than of severe illness and mortality among hospitalized patients. On the other hand, a number of studies have also found evidence that even in hospitalized populations, comorbidities such as hypertension, cardiovascular disease, CPD, and diabetes were predictive of severe illness or mortality [42,50-53]. However, even these findings are not necessarily inconsistent with our results since comorbidities and BMI tended to be “statistically significant” despite not meaningfully improving predictive performance. Furthermore, the wide range of mortality reported in case series of patients with severe COVID-19 may indicate that factors that predict severe disease do not necessarily predict mortality [54,55].

### Study Limitations

This study is not without limitations. First, there was considerable missing data, especially for laboratory results. We attempted to overcome this limitation using multiple imputation, although the coefficient estimates are only guaranteed to be unbiased if the data are missing at random and the missing mechanism is known. While this is an untestable assumption, our diagnostics were not suggestive of problems in the imputation as the distribution of the observed and imputed data were very similar.

Second, many of the laboratory results contained outliers. Although we truncated these variables to improve fit, predictions for new patients with extreme laboratory values lying outside of the chosen bounds are inherently uncertain. The presence of outliers could also imply that some laboratory values have been miscoded. This miscoding is a form of measurement error that would attenuate the relationship between mortality and the laboratory values [56,57].

Third, we did not have data on the day of death or out-of-hospital mortality. The latter could mean that mortality is underestimated if patients are discharged from the hospital and later die at home from COVID-19. Evidence suggests that COVID-19 deaths in the hospital comprise 38% of all deaths, but since the proportion of those 38% who were previously hospitalized is unknown, it is difficult to calibrate the extent of this potential bias [12]. Without day of death data, we were unable to perform time to event analyses that allowed for right censoring, raising the possibility of further underestimation of mortality. While we did aim to mitigate this limitation by dropping all observations with index dates less than 2 weeks from the data release date and controlling for calendar time in our models, future work should consider time to event analyses if the data permits.

## Conclusion

We developed a prognostic model of mortality with a cohort of over 17,000 patients hospitalized with COVID-19 in the U.S. using information on demographic, comorbidities, laboratory results and vital signs. We addressed many of the limitations of prior studies by using a large geographically diverse U.S. database, assessing calibration, performing validation using bootstrap resampling and a random training/test split, and providing detailed descriptions of the study population and statistical methods in the Supplement. Age was the strongest predictor of mortality and the predictive performance of a model that only included age was nearly identical to a model containing additional demographic information (age, sex, race/ethnicity, geographic location, smoking status, and calendar time) and a model containing information on both demographics and comorbidities. Vital signs and laboratory results did, however, add prognostic information beyond age. Overall, these results suggest that age, vital signs, and laboratory results may be useful to assess the prognosis of hospitalized patients, although external validation on new data is needed.

## Supporting information

Link to code and online supplement

## Data Availability

Data for this study was licensed from Optum. While we are unfortunately not able to publicly share the data, the Supplement (with all code and output for the manuscript as well as supplementary analyses) is available at https://github.com/phcanalytics/covid19-prognostic-model.

